# Effect of a novel food rich in miraculin on the oral microbiome of malnourished oncologic patients with dysgeusia

**DOI:** 10.1101/2024.07.12.24310343

**Authors:** Julio Plaza-Diaz, Francisco Javier Ruiz-Ojeda, Bricia López-Plaza, Marco Brandimonte-Hernández, Ana Isabel Álvarez-Mercado, Lucía Arcos-Castellanos, Jaime Feliú-Batlle, Thomas Hummel, Samara Palma-Milla, Angel Gil

## Abstract

Dysgeusia contributes to the derangement of nutritional status in patients with cancer, as well as worsening the quality of life. The pharmaceutical industry has failed to provide effective treatments for patients suffering from taste disorders. The present study provided a novel strategy to reduce side effects in patients with cancer through the administration of a novel food supplement approved by the European Union, Dried Miracle Berries (DMB), containing the taste-modifying glycoprotein miraculin, as an adjuvant to medical-nutritional treatment. This was done in a pilot randomized, parallel, triple-blind, and placebo-controlled intervention clinical trial in which 31 malnourished patients with cancer and dysgeusia receiving antineoplastic treatment were randomized into three arms [standard dose of DMB (150 mg DMB/tablet), high dose of DMB (300 mg DMB/tablet) or placebo (300 mg freeze-dried strawberry)] for three months. Patients consumed a DMB or placebo tablet before each main meal. Using the Nanopore methodology, we analyzed the oral microbiome of patients with cancer using saliva samples. All patients with cancer and dysgeusia had dysbiosis in terms of lower bacterial diversity and richness. DMB consumption was associated with changes in oral microbiome composition. Neither selected bacteria, nor taste perception, type of diet, and cytokine levels were associated with mucositis. Likewise, alcohol and tobacco consumption as well as general and digestive toxicity due to systemic therapy was not associated to specific changes of the oral microbiota. The standard dose of DMB resulted in a greater relative abundance of *Enterococcus* and a lower abundance of *Veillonella* compared with the high DMB dose and placebo. In particular, some species such as *Granulicatella elegans*, *Granulicatella adiacens*, *Streptococcus mutans*, and *Gemella morbillorum* showed higher relative abundances in the DMB standard-dose group; in contrast, *Streptococcus parasanguinis*, *Veillonella parvula*, *Streptococcus australis*, and *Streptococcus cristatus* were less abundant. Additionally, the consumption of a standard dose of DMB revealed a negative association between the concentrations of TNF-α and the abundance of species such as *Streptococcus thermophilus*, *Streptococcus pneumoniae*, *Streptococcus dysgalactiae* and *Streptococcus agalactiae.* Accordingly, regular DMB consumption changed the oral microbiome in patients with cancer and dysgeusia, which may contribute to maintaining an appropriate immune response without changing taste perception. However, as the present pilot study involved a small number of participants, further studies are necessary draw robust conclusions from the data.

**Highlights:**

- Patients with cancer and dysgeusia exhibit a dysbiotic state in terms of bacterial diversity and richness.
- The regular consumption of a standard dose of Dried Miracle Berries (DMB), rich in miraculin, before each main meal for three months as an adjuvant to medical-nutritional treatment, improves the oral microbiome composition in malnourished patients with cancer and dysgeusia.
- Several species i.e., *Granulicatella elegans*, *Granulicatella adiacens*, *Streptococcus mutans*, and *Gemella morbillorum*, show higher relative abundances in the DMB standard-dose group; in contrast, *Streptococcus parasanguinis*, *Veillonella parvula*, *Streptococcus australis*, and *Streptococcus cristatus* are less abundant
- DMB consumption is negatively associated with some species of *Streptococcus* and TNF-α concentrations in malnourished patients with cancer and dysgeusia.
- Neither of the highly represented bacteria are associated with the presence or absence of mucositis, digestive toxicity, or tobacco use and alcohol consumption or a change in taste perception at the end of the intervention.

## Introduction

Cancer is a group of diseases characterized by uncontrolled cell proliferation [1], affecting people in a multitude of ways, encompassing psychological, physiological, economic, and social aspects [2]. The 2020 data from the World Health Organization (WHO) indicate that 19.3 million new cancer cases diagnosed, and the number of rising cancer cases might reach 30.2 million by 2040. Hence, seeking treatments and prevention can reduce those costs, improve patients’ quality of life, and possibly increase survival rates.

Systemic cancer treatments such as chemotherapy, immunotherapy, and radiotherapy may lead to undesirable side effects that affect taste and smell [3, 4]. Indeed, patients with cancer undergoing systemic treatments often experience taste disorders in the range of 20-86% [5–7]. Different types of taste disorders have been reported in patients with cancer depending on the type of antineoplastic treatment, the location of the cancer and its characteristics [8, 9]. Dysgeusia is the most frequent qualitative and quantitative taste dysfunction, including taste distortions with bitter, metallic, salty, or unpleasant tastes [10, 11].

Additionally, antineoplastic therapies have also been associated with reduced saliva production [12–15]. Thus, xerostomia may also cause the saliva to be thicker and contain high concentrations of salt, influencing the sense of taste [12, 16].

A reduction in quality of life in patients with cancer is observed when taste is impaired with further deterioration of their nutritional status [17]. In addition to reducing the patient’s quality of life, dysgeusia can lead to weight loss during treatment and worsen the patient’s prognosis [18]. The pharmaceutical industry has failed to provide effective treatments for dysgeusia. The treatments used for taste disorders are zinc supplementation, amifostine, selenium, lactoferrin, and cannabinoids; however, these treatments exhibit limited efficacy [19, 20]. Indeed, to reduce dysgeusia and improve patient health and quality of life, novel strategies are needed [21, 22].

The oral microbiota includes protozoans, fungi, yeast, bacteria, archaea, viruses and phages [23, 24], with bacteria being the most studied element [25]. Many factors affect individual oral microbiota, including age, diet, and lifestyle habits (*e.g*., smoking and alcohol consumption) [26, 27]. Moreover, the oral microbiota plays a significant role in health, especially in terms of the oral cavity. Indeed, this microbial community is correlated with several oral pathologies, such as oral and other types of cancer [24]. The majority of studies showing an association between derangement of the oral microbiota and oral diseases induced by cancer therapies were performed in patients undergoing radiotherapy [28, 29]. Hence, a reduction in diversity and richness seems to be associated with the development of oral diseases [30].

Over the past decade, there has been a significant increase in research into the prevention and treatment of oral diseases [31]. This includes the identification of bioactive food components and the development of functional foods with health-promoting benefits [32]. Indeed, it is possible to increase patients’ quality of life with the supplementation of functional foods. Consequently, concerning taste disorders, oncologic patients may switch from traditional therapies to innovative alternatives that can have a positive outcome [33].

Miraculin is a glycoprotein that converts sour flavors into sweet ones and increases enjoyment of meals [34, 35]. This protein is present in *Synsepalum dulcificum*, a plant native to West Africa that is commonly known as the “miracle berry”. The taste-modifying effect of this miracle fruit is effective under acidic conditions and lasts for approximately 30-60 minutes [36]. The freeze-dried extract of the miracle berries pulp, rich in miraculin, is called dried miracle berries (DMB) and was approved by the European Commission as a Novel Food in December 2021 [37]. DMB is not only interesting because of its taste-modifying effects but also because of its bioactive ingredients, including fiber and phenolic compounds [38, 39].

Only two small trials have reported the potential of this berry for improving dysgeusia caused by chemotherapy [18, 40]. Recently, the research group generated clinical evidence on the efficacy of the DMB for the management of dysgeusia in a pilot randomized, parallel, triple-blind, and placebo-controlled clinical trial (the CLINMIR study) [41, 42]. In that study, we observed improvements in electrochemical food perception, energy and nutrient intake, nutritional status, and quality of life in malnourished patients with cancer receiving antineoplastic treatment [41]. In this paper, our objective was to assess the oral microbiome of malnourished patients with cancer and dysgeusia after DMB consumption as a medical-nutritional adjuvant treatment.

## Material and methods

### Ethical statement

Scientific Research and Ethics Committee approval was obtained in June 2022 from University Hospital La Paz (HULP, Code 6164). This study adheres to the Ethical Standards of the Declaration of Helsinki concerning recommendations that guide physicians conducting biomedical research on humans. All researchers should become familiar with and follow the ICH Harmonized Tripartite Guidelines to follow good clinical practice [43].

Participants signed the informed consent form; researchers informed them of the study characteristics and what participation in the trial entailed (verbally and in writing). We followed several legal requirements when processing personal information, including the Spanish Organic Law 3/2018 of 5 December and the General Data Protection Regulation of the European Union (EU) 2016/679 of 27 April 2016.

### Experimental design and participants

The CLINMIR study is a pilot randomized, parallel, triple-blind, and placebo-controlled clinical trial. Using the number NCT05486260, the present protocol was registered at http://clinicaltrials.gov, accessed on 14 March 2024. The oncology service and the clinical nutrition unit at HULP in Madrid recruited 31 malnourished patients with cancer and dysgeusia [42].

Patients over 18 years of age who were treated for cancer using chemotherapy, radiotherapy, and/or immunotherapy and had lost 5% of their weight, were defined as malnourished according to GLIM criteria [44], and those who suffered dysgeusia, measured by electrogustometry, were included in the study. Additionally, these patients should have a three-month life expectancy [41].

The study excluded participants who were involved in another clinical trial, received enteral or parenteral nutrition or had poorly controlled diabetes mellitus (HbA1c > 8%), hypertension or uncontrolled hyperthyroidism, severe digestive toxicity as a result of chemotherapy and radiotherapy, or who were suffering from severe kidney or liver disease (chronic renal failure, nephrotic syndrome, cirrhosis, etc.). Additionally, participants should not suffer from severe dementia, brain metastases, eating disorders, a history of severe neurological or psychiatric disorders that may interfere with treatment, alcoholism or substance abuse, or severe gastrointestinal diseases.

Malnourished patients with cancer and dysgeusia who were receiving active treatment were randomly assigned to one of three treatment arms. In a three-month study, each patient was instructed to dissolve a miraculin-based food supplement tablet five minutes before each main meal (breakfast, lunch, and dinner). Patients meeting the selection criteria were randomly assigned to one of the three groups in the clinical trial. The first group received a standard dose of DMB (150 mg DMB, equivalent to 2.8 mg of miraculin + 150 mg freeze-dried strawberry per orodispersible tablet). The second group had a higher dose of DMB (300 mg DMB, equivalent to 5.6 mg of miraculin). Finally, the third group took a placebo dose (300 mg of freeze-dried strawberry). All three treatments were isocaloric (Table S1). The clinical trial consisted of two phases. There were six face-to-face meetings in each. The selection phase had one selection visit, whereas the experimental phase had five visits. All participants were undergoing active treatment with at least chemotherapy, and taste alterations were measured by electrogustometry and taste strip tests [42]. To complete the three-month intervention period, the subjects received as many tablets as necessary during scheduled visits to the HULP. Participants were asked to return all packaging, regardless of whether it was empty or partially consumed. This was done to assess compliance by comparing the number of tablets provided and returned. DMB was administered as an adjuvant medical-nutritional treatment [41, 42, 45].

### Biological samples and DNA sequencing

The trial lasted for three months. For each intervention group (standard dose, high dose, and placebo), saliva samples were analyzed for the oral microbiome. Before the analysis, saliva samples were collected at baseline, 1 month after treatment, and 3 months after intervention, and placed in OMNIgene Oral OM-501 tubes (DNA Genotek Inc., Ottawa, ON, Canada).

To avoid more punctures and hospital visits than necessary, blood samples were collected by trained personnel at the HULP Extraction Unit in the morning (approximately 8:00 am) during blood tests before chemotherapy. Blood samples were collected in vacuum tubes, labeled, transported, and centrifuged for 10 minutes at 1500 x g. Aliquots of blood samples were prepared and labeled according to a numerical code and stored at -80°C [41].

### Saliva DNA extraction

DNA was extracted from saliva samples using QIAamp DNA Microbiome Kit (Ref. ID: 51704, Qiagen Inc., Hilden, Germany) according to manufacturer instructions. DNA purity and integrity were assessed using spectrophotometry (NanoDrop, Thermo Fisher Scientific, Massachusetts, USA).

### Whole 16S rRNA gene sequencing and taxonomic assignment

Following the instructions of the 16S Barcoding kit ref SQK-16S024 (Oxford Nanopore Technologies, Oxford, United Kingdom), the 16S rRNA gene was PCR-amplified using redesigned 16S primers (27F and 1492R) with 5’ tags that facilitate the ligase-free attachment of Rapid Sequencing Adapters. A 0.2 ml PCR tube was filled with RNA-free water, 14 μl; 10 ng of input DNA, 10 ml; 16S barcodes at 10 ml, 1 μl; and LongAmp Taq 2X Master mix, 25 μl, for a total volume of 50 μl per sample. The following conditions were used for PCR: Denaturation at 95 °C for 1 minute, 25 cycles of denaturation at 95 °C for 20 seconds, banding at 55 °C for 30 seconds, and extension at 65 °C for 2 minutes, followed by a final extension at 65 °C for 5 minutes.

A new 1.5 ml Eppendorf DNA LoBind tube was used to transfer the PCR product. By vortexing 30 ml of AMPure XP beads and mixing by pipetting, PCR products from each sample were resuspended in AMPure XP beads (Beckman Coulter, ThermoScientific, Spain), and cleaned in 10 μl of 10 mM Tris-HCl pH 8.0 with 50 mM NaCl. Finally, all barcoded libraries from each sample were combined in the appropriate ratios to obtain a total concentration of 50-100 fmoles. The 16S amplicon of 1500 bp corresponds to those 50-100 fmoles. In the final step, 1 μl of rapid adapter tube was added to the previously mixed barcoded DNA with which each sample was identified. The mixture should be mixed gently by shaking the tube and centrifuged. A final volume of 11 μl is obtained by incubating the reaction for 5 minutes at room temperature.

A fresh tube of the library was prepared for loading into SpotON Flow Cell Mk R9 Version (ref FLO-MIN106D, Oxford Nanopore Technologies, Oxford, United Kingdom) using the Minion M1kc and M1kb sequencers (Oxford Nanopore Technologies, Oxford, United Kingdom). A total of 75 μl was used for the following components: sequencing buffer (SQB) 34 μl, loading beads (LB), mixed immediately before use 25.5 μl, RNA-free water 4.5 μl, and a previously prepared DNA library incubated at room temperature 11 μl.

Once the raw data were generated, the base calling was performed with Guppy version 6.5.7 (Oxford Nanopore Technologies, Oxford, United Kingdom), and the resulting sequences were identified using Kraken2 (with Refseq Archaea, bacteria, viral, plasmid, human, UniVec_Core, protozoa, fungi & plant database) and further analyzed using QIIME (2-2020.8) [46]. The taxonomy was assigned to ASVs using the sklearn naïve Bayes taxonomy classifier (via q2-feature-classifier) [47] against SILVA 16S V3-V4 v132_99 [48] with a similarity threshold of 99%. Phylum, family, genus, and species levels were used for the interpretation of the results. Using the vegan library [49], the Shannon and Simpson’s indices were used to examine the diversity of the samples, and the Chao1 index was used to estimate species richness.

### Plasma cytokines

Tumor necrosis factor-alpha (TNF-α), and human proteolysis-inducing factor/dermcidin (PIF) were determined and analyzed as previously described [41, 42, 45].

### Electrical taste perception

Electric-induced taste stimulus (taste acuity) was measured by electrogustometry as described previously [41, 45].

### Dietary pattern assessment

Food daily records were recorded for three days, one of which was a holiday (weekend day, a day off, or a day out of the usual routine). In the absence of weight recording, patients were advised to record household measurements (spoonful, cups, etc.) or to record household weights. During the review of all records, a nutritionist ensured that all the information collected was accurate and complete in the presence of the patient. With the help of DIAL software (Alce Ingeniera, Madrid, Spain), the energy and nutrients contained in foods, drinks, dietary supplements, and preparations were converted into energy and nutrients. The results were compared with the recommended intakes for the Spanish population [41].

### Mucositis

At baseline, participants were scheduled for appointments. Participants underwent an oral examination with the use of mouth mirrors and a high-power headlamp at each appointment (5 visits) by a previously-calibrated investigator. Oral mucositis clinical signs were recorded using the clinical component of the National Cancer Institute Common Terminology Criteria for Adverse Events version 3 (NCI-CTCAE v. 3) [50] and the clinical component of the Oral Mucositis Assessment Scale during the course of the intervention [51].

### Statistical analysis

A linear mixed model was used to evaluate the differences between placebo, DMB 150 mg, and DMB 300 mg, as well as the differences between visits: the effects of treatment, time, and their interaction (time x treatment) were considered. The linear mixed model was developed using the lme4 package [52] in the R program [53]. Further, general linear mixed models (GLM) of covariance (ANCOVA) were used to evaluate differences between means for treatment, time, and treatment x time using baseline data as covariates (SPSS Inc., Chicago, IL, USA). Post-hoc analyses were determined using the Bonferroni test. Compared to other programs, the lme4 package may be faster and more memory-efficient because it employs modern, efficient linear algebra methods, such as those in the Eigen package, and reference classes to prevent undue copying of large objects. Moreover, the software provides the ability to construct generalized linear mixed models, which maximizes the amount of information available when loss patients are included in some of the analyzed conditions [52].

This study examined the relationships between oral microbiome variables, inflammatory parameters, dietary variables, and electrical taste perception outcomes using Pearson’s correlations. R Studio’s corrplot function [54] was used to express associations by correcting multiple tests with the false discovery rate (FDR) procedure [55]. The graphs show only significant and corrected associations. In the graphs, red and blue lines indicate the correlation values, with negative correlations highlighted in red (−1) and positive correlations highlighted in blue (+1).

Based on a Rivera-Pinto analysis, it is possible to identify microbial signatures, i.e., groups of microbes that can predict particular phenotypes of interest [56]. These microbial signatures correspond to an individual’s unique microbiome and may be used to diagnose, prognosticate, or predict therapeutic responses. To identify microbial signatures, we can model the response variable as well as select the taxa that yield the highest level of classification or prediction accuracy. As part of the Rivera-Pinto method and the Selbal algorithm, we evaluated specific signatures at the phylum and genus levels to select a sparse model that adequately explains the response variable. The microbial signatures were calculated using geometric means based on data collected from two groups of taxa. The name implies that these groups are those with relative abundances, or balances, that are related to the response variable of interest [56].

Finally, it was examined whether the presence or appearance of mucositis could be independently correlated with the response to the DMB treatment, the levels of cytokine, taste perception, dietary parameters, and the presence of selected species. Additionally, we examined clinical outcomes in order to determine whether they could be independently correlated with the presence of specific bacterial species. Our study evaluated the presence or absence of immunotherapeutic agents (including atezolizumad, becacizumab, paclitaxel, pembrolizumab, nivolumab, panitumumab, folfirinox, among others), general toxicity (including neutropenia, diarrhea, thrombocytopenia, among others), digestive toxicity, tobacco use and alcohol consumption. To achieve this, we used the SPSS statistical package (SPSS Inc, Chicago, IL) to perform a binary logistic regression with the Wald regression backward option and a significance level of p<0.05.

## Results

Patients were recruited from November 2022 to May 2023 and 62 were assessed for eligibility. 31 patients with cancer and dysgeusia were included in the study for meeting the selection criteria and were randomly assigned to one of three intervention groups, which were adjusted according to the type of cancer. During the follow-up period, which extended from November 2022 to August 2023, there were 10 instances of participants dropping out of the study. The majority of these drop-outs were due to the taste distortion of non-sweet acidic foods (n = 6) and the difficulty that the prescription derived from the intervention added to their already complex antineoplastic treatment (n = 2). Furthermore, two placebo patients died. Finally, a total of 21 oncological patients completed the clinical trial. All variables were evaluated following the intention-to-treat principle [42]. The sample comprised 58.1% women and 41.9% men, with a mean age of 60.0 ± 10.9 years. Baseline data for the population have been published previously elsewhere [41].

### Phylum and family levels

In all groups, the relative abundances at the phylum level were similar at baseline, after one month, and after three months. *Bacillota* accounted for more than 98% of the relative abundance of the oral microbiome in our study. As for the other main phyla, *Actinobacteriota*, *Fusobacteria*, *Bacteroidota*, *Pseudomonadota*, and *Saccharibacteria*, there were no significant changes or trends among the groups (**Table S2**). According to Shannon and Simpson indices, related to bacterial diversity, there was no significant difference between the groups. Additionally, there were no differences among the groups in terms of the Chao1 index, which is related to bacterial species richness. The patients with cancer and dysgeusia showed dysbiosis in terms of diversity and richness, compared with healthy individuals [57]. At the family level, we did not observe significant changes in the oral microbiome of patients with cancer either during treatment or time (**Table S3**).

### Genus level

At the genus level, *Streptococcus* was the most common in all studied groups (more than 63%). We observed significant differences in the interaction between treatment and time for *Enterococcus* and *Veillonella*. The consumption of the standard dose of DMB resulted in a significantly greater relative abundance of *Enterococcus* than in the high dose of DMB and placebo groups (**Table 1**). *Veillonella* genus relative abundance decreased significantly over the treatment period in the standard dose DMB group compared to the high dose DMB and placebo groups (**Table 1**). In addition, we observed a trend of increase for *Granulicatella* (p=0.066)*, Bacillus* (p=0.061), and *Staphylococcus* (p=0.053) in DMB groups (**Table 1**). The remaining genera did not show any significant changes, in either the treatment or time.

**Table 1.**
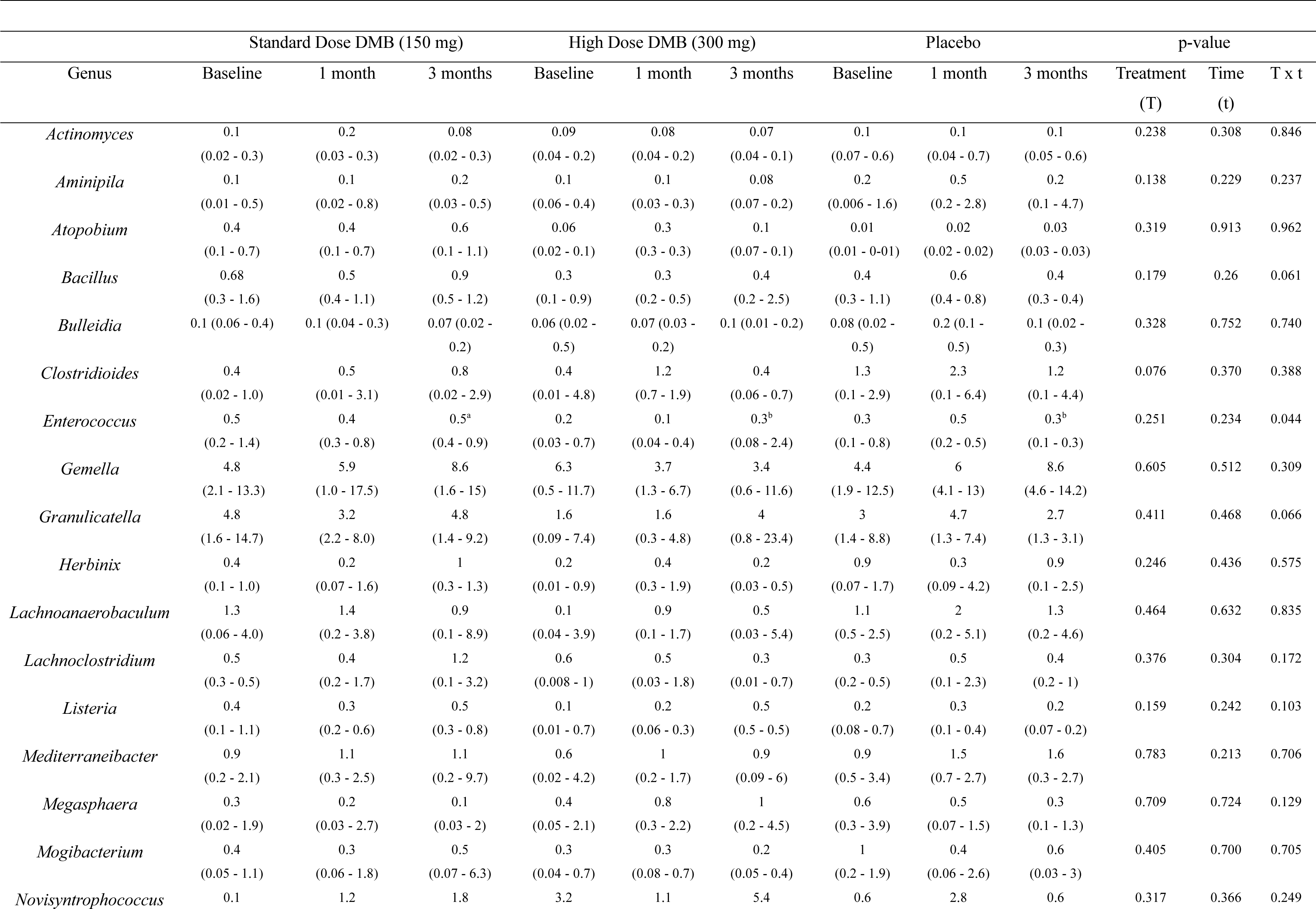
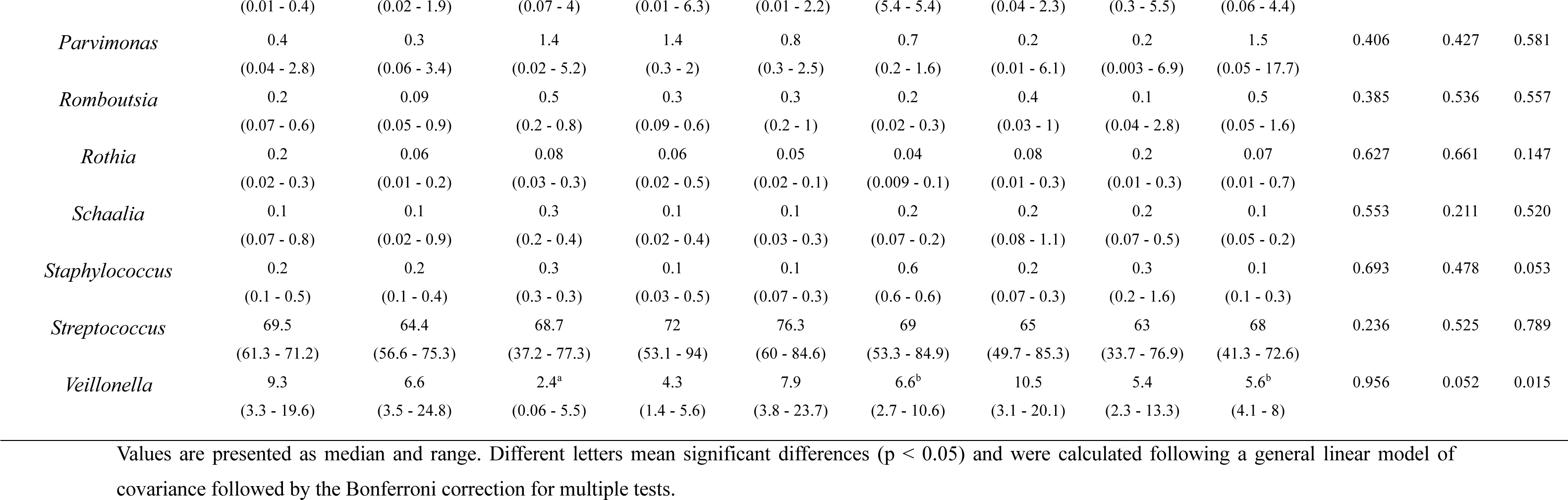
Relative abundances at the genus levels of oral bacteria in malnourished patients with cancer and dysgeusia who received the standard dose of DMB (150 mg/tablet), the high dose of DMB(300 mg/tablet) or the placebo for 3 months.

| Genus | Standard Dose DMB (150 mg) |  |  | High Dose DMB (300 mg) |  |  | Placebo |  |  | p-value |  |  |
| --- | --- | --- | --- | --- | --- | --- | --- | --- | --- | --- | --- | --- |
|  | Baseline | 1 month | 3 months | Baseline | 1 month | 3 months | Baseline | 1 month | 3 months | Treatment<br>(T) | Time<br>(t) | T x t |
| *Actinomyces* | 0.1<br>(0.02 - 0.3) | 0.2<br>(0.03 - 0.3) | 0.08<br>(0.02 - 0.3) | 0.09<br>(0.04 - 0.2) | 0.08<br>(0.04 - 0.2) | 0.07<br>(0.04 - 0.1) | 0.1<br>(0.07 - 0.6) | 0.1<br>(0.04 - 0.7) | 0.1<br>(0.05 - 0.6) | 0.238 | 0.308 | 0.846 |
| *Aminipila* | 0.1<br>(0.01 - 0.5) | 0.1<br>(0.02 - 0.8) | 0.2<br>(0.03 - 0.5) | 0.1<br>(0.06 - 0.4) | 0.1<br>(0.03 - 0.3) | 0.08<br>(0.07 - 0.2) | 0.2<br>(0.006 - 1.6) | 0.5<br>(0.2 - 2.8) | 0.2<br>(0.1 - 4.7) | 0.138 | 0.229 | 0.237 |
| *Atopobium* | 0.4<br>(0.1 - 0.7) | 0.4<br>(0.1 - 0.7) | 0.6<br>(0.1 - 1.1) | 0.06<br>(0.02 - 0.1) | 0.3<br>(0.3 - 0.3) | 0.1<br>(0.07 - 0.1) | 0.01<br>(0.01 - 0.01) | 0.02<br>(0.02 - 0.02) | 0.03<br>(0.03 - 0.03) | 0.319 | 0.913 | 0.962 |
| *Bacillus* | 0.68<br>(0.3 - 1.6) | 0.5<br>(0.4 - 1.1) | 0.9<br>(0.5 - 1.2) | 0.3<br>(0.1 - 0.9) | 0.3<br>(0.2 - 0.5) | 0.4<br>(0.2 - 2.5) | 0.4<br>(0.3 - 1.1) | 0.6<br>(0.4 - 0.8) | 0.4<br>(0.3 - 0.4) | 0.179 | 0.26 | 0.061 |
| *Bulleidia* | 0.1 (0.06 - 0.4) | 0.1 (0.04 - 0.3) | 0.07 (0.02 - 0.2) | 0.06 (0.02 - 0.5) | 0.07 (0.03 - 0.2) | 0.1 (0.01 - 0.2) | 0.08 (0.02 - 0.5) | 0.2 (0.1 - 0.5) | 0.1 (0.02 - 0.3) | 0.328 | 0.752 | 0.740 |
| *Clostridioides* | 0.4<br>(0.02 - 1.0) | 0.5<br>(0.01 - 3.1) | 0.8<br>(0.02 - 2.9) | 0.4<br>(0.01 - 4.8) | 1.2<br>(0.7 - 1.9) | 0.4<br>(0.06 - 0.7) | 1.3<br>(0.1 - 2.9) | 2.3<br>(0.1 - 6.4) | 1.2<br>(0.1 - 4.4) | 0.076 | 0.370 | 0.388 |
| *Enterococcus* | 0.5<br>(0.2 - 1.4) | 0.4<br>(0.3 - 0.8) | 0.5<sup>a</sup><br>(0.4 - 0.9) | 0.2<br>(0.03 - 0.7) | 0.1<br>(0.04 - 0.4) | 0.3<sup>b</sup><br>(0.08 - 2.4) | 0.3<br>(0.1 - 0.8) | 0.5<br>(0.2 - 0.5) | 0.3<sup>b</sup><br>(0.1 - 0.3) | 0.251 | 0.234 | 0.044 |
| *Gemella* | 4.8<br>(2.1 - 13.3) | 5.9<br>(1.0 - 17.5) | 8.6<br>(1.6 - 15) | 6.3<br>(0.5 - 11.7) | 3.7<br>(1.3 - 6.7) | 3.4<br>(0.6 - 11.6) | 4.4<br>(1.9 - 12.5) | 6<br>(4.1 - 13) | 8.6<br>(4.6 - 14.2) | 0.605 | 0.512 | 0.309 |
| *Granulicatella* | 4.8<br>(1.6 - 14.7) | 3.2<br>(2.2 - 8.0) | 4.8<br>(1.4 - 9.2) | 1.6<br>(0.09 - 7.4) | 1.6<br>(0.3 - 4.8) | 4<br>(0.8 - 23.4) | 3<br>(1.4 - 8.8) | 4.7<br>(1.3 - 7.4) | 2.7<br>(1.3 - 3.1) | 0.411 | 0.468 | 0.066 |
| *Herbinix* | 0.4<br>(0.1 - 1.0) | 0.2<br>(0.07 - 1.6) | 1<br>(0.3 - 1.3) | 0.2<br>(0.01 - 0.9) | 0.4<br>(0.3 - 1.9) | 0.2<br>(0.03 - 0.5) | 0.9<br>(0.07 - 1.7) | 0.3<br>(0.09 - 4.2) | 0.9<br>(0.1 - 2.5) | 0.246 | 0.436 | 0.575 |
| *Lachnoanaerobaculum* | 1.3<br>(0.06 - 4.0) | 1.4<br>(0.2 - 3.8) | 0.9<br>(0.1 - 8.9) | 0.1<br>(0.04 - 3.9) | 0.9<br>(0.1 - 1.7) | 0.5<br>(0.03 - 5.4) | 1.1<br>(0.5 - 2.5) | 2<br>(0.2 - 5.1) | 1.3<br>(0.2 - 4.6) | 0.464 | 0.632 | 0.835 |
| *Lachnoclostridium* | 0.5<br>(0.3 - 0.5) | 0.4<br>(0.2 - 1.7) | 1.2<br>(0.1 - 3.2) | 0.6<br>(0.008 - 1) | 0.5<br>(0.03 - 1.8) | 0.3<br>(0.01 - 0.7) | 0.3<br>(0.2 - 0.5) | 0.5<br>(0.1 - 2.3) | 0.4<br>(0.2 - 1) | 0.376 | 0.304 | 0.172 |
| *Listeria* | 0.4<br>(0.1 - 1.1) | 0.3<br>(0.2 - 0.6) | 0.5<br>(0.3 - 0.8) | 0.1<br>(0.01 - 0.7) | 0.2<br>(0.06 - 0.3) | 0.5<br>(0.5 - 0.5) | 0.2<br>(0.08 - 0.7) | 0.3<br>(0.1 - 0.4) | 0.2<br>(0.07 - 0.2) | 0.159 | 0.242 | 0.103 |
| *Mediterraneibacter* | 0.9<br>(0.2 - 2.1) | 1.1<br>(0.3 - 2.5) | 1.1<br>(0.2 - 9.7) | 0.6<br>(0.02 - 4.2) | 1<br>(0.2 - 1.7) | 0.9<br>(0.09 - 6) | 0.9<br>(0.5 - 3.4) | 1.5<br>(0.7 - 2.7) | 1.6<br>(0.3 - 2.7) | 0.783 | 0.213 | 0.706 |
| *Megasphaera* | 0.3<br>(0.02 - 1.9) | 0.2<br>(0.03 - 2.7) | 0.1<br>(0.03 - 2) | 0.4<br>(0.05 - 2.1) | 0.8<br>(0.3 - 2.2) | 1<br>(0.2 - 4.5) | 0.6<br>(0.3 - 3.9) | 0.5<br>(0.07 - 1.5) | 0.3<br>(0.1 - 1.3) | 0.709 | 0.724 | 0.129 |
| *Mogibacterium* | 0.4<br>(0.05 - 1.1) | 0.3<br>(0.06 - 1.8) | 0.5<br>(0.07 - 6.3) | 0.3<br>(0.04 - 0.7) | 0.3<br>(0.08 - 0.7) | 0.2<br>(0.05 - 0.4) | 1<br>(0.2 - 1.9) | 0.4<br>(0.06 - 2.6) | 0.6<br>(0.03 - 3) | 0.405 | 0.700 | 0.705 |
| *Novisyntrophococcus* | 0.1 | 1.2 | 1.8 | 3.2 | 1.1 | 5.4 | 0.6 | 2.8 | 0.6 | 0.317 | 0.366 | 0.249 |
|  | (0.01 - 0.4) | (0.02 - 1.9) | (0.07 - 4) | (0.01 - 6.3) | (0.01 - 2.2) | (5.4 - 5.4) | (0.04 - 2.3) | (0.3 - 5.5) | (0.06 - 4.4) |  |  |  |
| *Parvimonas* | 0.4 | 0.3 | 1.4 | 1.4 | 0.8 | 0.7 | 0.2 | 0.2 | 1.5 | 0.406 | 0.427 | 0.581 |
|  | (0.04 - 2.8) | (0.06 - 3.4) | (0.02 - 5.2) | (0.3 - 2) | (0.3 - 2.5) | (0.2 - 1.6) | (0.01 - 6.1) | (0.003 - 6.9) | (0.05 - 17.7) |  |  |  |
| *Romboutsia* | 0.2 | 0.09 | 0.5 | 0.3 | 0.3 | 0.2 | 0.4 | 0.1 | 0.5 | 0.385 | 0.536 | 0.557 |
|  | (0.07 - 0.6) | (0.05 - 0.9) | (0.2 - 0.8) | (0.09 - 0.6) | (0.2 - 1) | (0.02 - 0.3) | (0.03 - 1) | (0.04 - 2.8) | (0.05 - 1.6) |  |  |  |
| *Rothia* | 0.2 | 0.06 | 0.08 | 0.06 | 0.05 | 0.04 | 0.08 | 0.2 | 0.07 | 0.627 | 0.661 | 0.147 |
|  | (0.02 - 0.3) | (0.01 - 0.2) | (0.03 - 0.3) | (0.02 - 0.5) | (0.02 - 0.1) | (0.009 - 0.1) | (0.01 - 0.3) | (0.01 - 0.3) | (0.01 - 0.7) |  |  |  |
| *Schaalia* | 0.1 | 0.1 | 0.3 | 0.1 | 0.1 | 0.2 | 0.2 | 0.2 | 0.1 | 0.553 | 0.211 | 0.520 |
|  | (0.07 - 0.8) | (0.02 - 0.9) | (0.2 - 0.4) | (0.02 - 0.4) | (0.03 - 0.3) | (0.07 - 0.2) | (0.08 - 1.1) | (0.07 - 0.5) | (0.05 - 0.2) |  |  |  |
| *Staphylococcus* | 0.2 | 0.2 | 0.3 | 0.1 | 0.1 | 0.6 | 0.2 | 0.3 | 0.1 | 0.693 | 0.478 | 0.053 |
|  | (0.1 - 0.5) | (0.1 - 0.4) | (0.3 - 0.3) | (0.03 - 0.5) | (0.07 - 0.3) | (0.6 - 0.6) | (0.07 - 0.3) | (0.2 - 1.6) | (0.1 - 0.3) |  |  |  |
| *Streptococcus* | 69.5 | 64.4 | 68.7 | 72 | 76.3 | 69 | 65 | 63 | 68 | 0.236 | 0.525 | 0.789 |
|  | (61.3 - 71.2) | (56.6 - 75.3) | (37.2 - 77.3) | (53.1 - 94) | (60 - 84.6) | (53.3 - 84.9) | (49.7 - 85.3) | (33.7 - 76.9) | (41.3 - 72.6) |  |  |  |
| *Veillonella* | 9.3 | 6.6 | 2.4<sup>a</sup> | 4.3 | 7.9 | 6.6<sup>b</sup> | 10.5 | 5.4 | 5.6<sup>b</sup> | 0.956 | 0.052 | 0.015 |
|  | (3.3 - 19.6) | (3.5 - 24.8) | (0.06 - 5.5) | (1.4 - 5.6) | (3.8 - 23.7) | (2.7 - 10.6) | (3.1 - 20.1) | (2.3 - 13.3) | (4.1 - 8) |  |  |  |

Values are presented as median and range. Different letters mean significant differences (p < 0.05) and were calculated following a general linear model of covariance followed by the Bonferroni correction for multiple tests.

### Species level

Three species dominated the oral microbiome of patients with cancer, *Streptococcus pneumoniae*, *Streptococcus thermophilus* and *Veillonella parvula*. We observed significant differences in the interaction between treatment and time for *Granulicatella elegans, Streptococcus mutans*, *Streptococcus parasanguinis* and *Veillonella parvula*, as well as a trend for *Gemella morbillorum* (p=0.054)*, Granulicatella adiacens* (p=0.069)*, Streptococcus australis* (p=0.063), and *Streptococcus cristatus* (p=0.087) (**Table 2**).

**Table 2.** Relative abundances of species for oral bacteria in malnourished patients with cancer and dysgeusia who received the standard dose of DMB (150 mg), the high dose of DMB (300 mg) or the placebo for 3 months.

|  | Species |  |  |  |  |  |  |  |  |  |  |  |
| --- | --- | --- | --- | --- | --- | --- | --- | --- | --- | --- | --- | --- |
|  | Standard Dose DMB (150 mg) |  |  | High Dose DMB (300 mg) |  |  | Placebo |  |  | p-value |  |  |
|  | Baseline | 1 month | 3 months | Baseline | 1 month | 3 months | Baseline | 1 month | 3 months | Treatment<br>(T) | Time (t) | T x t |
| *Gemella haemolysans* | 0.3<br>(0.06 - 1.4) | 0.2<br>(0.03 - 0.6) | 0.3<br>(0.07 - 1.2) | 0.6<br>(0.03 - 7.6) | 0.5<br>(0.2 - 0.7) | 0.5<br>(0.03 - 2.7) | 0.7<br>(0.1 - 6.1) | 0.7<br>(0.1 - 2.2) | 1.8<br>(0.3 - 6.7) | 0.105 | 0.297 | 0.517 |
| *Gemella morbillorum* | 0.1<br>(0.06 - 0.9) | 0.2<br>(0.07 - 0.6) | 0.3<br>(0.07 - 1.2) | 0.4<br>(0.04 - 2.2) | 0.4<br>(0.03 - 1.7) | 0.08<br>(0.06 - 0.5) | 0.3<br>(0.03 - 3.9) | 0.6<br>(0.08 - 9.4) | 2.5<br>(0.2 - 7.2) | 0.103 | 0.170 | 0.054 |
| *Gemella sanguinis* | 3.7<br>(1.4 - 12.8) | 5.3<br>(0.6 - 16.8) | 7.5<br>(1.3 - 15.1) | 1.0<br>(0.009 - 8.0) | 1.9<br>(0.1 - 6.0) | 1.1<br>(0.06 - 9.7) | 2.2<br>(1.2 - 6.1) | 3.2<br>(1.8 - 6.2) | 4.0<br>(0.4 - 7.0) | 0.312 | 0.352 | 0.769 |
| *Gemella sp. zg-570* | 0.06<br>(0.01 - 0.5) | 0.06<br>(0.02 - 0.2) | 0.1<br>(0.04 - 0.3) | 0.3<br>(0.04 - 1.8) | 0.3<br>(0.2 - 0.4) | 0.4<br>(0.01 - 0.6) | 0.2<br>(0.04 - 1.3) | 0.4<br>(0.06 - 0.5) | 0.4<br>(0.06 - 0.8) | 0.102 | 0.300 | 0.584 |
| *Granulicatella adiacens* | 4.8<br>(1.6 - 14.7) | 3.7<br>(2.2 - 8.0) | 4.8<br>(1.4 - 9.2) | 1.4<br>(0.09 - 7.4) | 1.6<br>(0.3 - 4.1) | 4.0<br>(0.8 - 23.6) | 3.0<br>(1.4 - 8.8) | 4.7<br>(1.3 - 7.5) | 2.6<br>(1.3 - 3.1) | 0.396 | 0.484 | 0.069 |
| *Granulicatella elegans* | 0.01<br>(0.004 - 0.1) | 0.02<br>(0.01 - 0.1) | 0.03<sup>a</sup><br>(0.01 - 0.07) | 0.02<br>(0.009 - 0.5) | 0.6<br>(0.6 - 0.6) | 0.01<sup>b</sup><br>(0.006 - 0.02) | 0.03<br>(0.01 - 0.2) | 0.01<br>(0.01 - 0.3) | 0.02<sup>b</sup><br>(0.005 - 0.05) | <0.001 | <0.001 | <0.001 |
| *Streptococcus agalactiae* | 10.5<br>(5.1 - 15.4) | 10.0<br>(5.6 - 14.2) | 9.8<br>(6.0 - 13.9) | 10.9<br>(5.5 - 14.1) | 7.8<br>(6.6 - 11.9) | 8.0<br>(5.9 - 11.4) | 9.2<br>(7.3 - 12.6) | 8.7<br>(6.2 - 14.2) | 7.9<br>(7.3 - 16.3) | 0.878 | 0.385 | 0.586 |
| *Streptococcus australis* | 0.1<br>(0.01 - 0.7) | 0.07<br>(0.02 - 0.2) | 0.06<br>(0.006 - 0.2) | 0.03<br>(0.02 - 0.03) | 0.03<br>(0.03 - 0.03) | 0.06<br>(0.06 - 0.06) | 0.01<br>(0.01 - 0.07) | 0<br>(0 - 0) | 0.6<br>(0.6 - 0.6) | 0.218 | 0.301 | 0.063 |
| *Streptococcus cristatus* | 0.4<br>(0.04 - 1.6) | 0.3<br>(0.06 - 1.5) | 0.1<br>(0.03 - 3.0) | 0.4<br>(0.1 - 4.0) | 1.2<br>(0.1 - 1.5) | 0.2<br>(0.04 - 0.7) | 0.7<br>(0.06 - 3.4) | 0.4<br>(0.06 - 0.7) | 0.3<br>(0.05 - 2.2) | 0.733 | 0.149 | 0.087 |
| *Streptococcus dysgalactiae* | 5.0<br>(3.3 - 6.3) | 4.8<br>(2.9 - 7.5) | 4.2<br>(2.5 - 5.8) | 4.5<br>(3.3 - 5.6) | 3.8<br>(3.3 - 6.0) | 3.6<br>(2.7 - 5.3) | 3.9<br>(2.8 - 6.2) | 3.6<br>(3.1 - 5.5) | 3.9<br>(3.1 - 6.3) | 0.583 | 0.261 | 0.666 |
| *Streptococcus infantis* | 1.0<br>(0.7 - 2.4) | 1.0<br>(0.6 - 2.8) | 0.9<br>(0.4 - 2.3) | 0.9<br>(0.5 - 1.2) | 0.7<br>(0.5 - 1.3) | 0.7<br>(0.6 - 1.0) | 0.9<br>(0.7 - 1.6) | 0.9<br>(0.6 - 1.7) | 1.1<br>(0.7 - 1.2) | 0.321 | 0.844 | 0.307 |
| *Streptococcus mutans* | 0.3<br>(0.2 - 2.9) | 0.3<br>(0.2 - 3.5) | 0.4<sup>a</sup><br>(0.2 - 3.4) | 1.9<br>(0.3 - 2.7) | 2.2<br>(0.4 - 7.2) | 0.6<sup>b</sup><br>(0.3 - 1.8) | 0.4<br>(0.2 - 1.2) | 0.3<br>(0.2 - 0.3) | 0.6<sup>b</sup><br>(0.2 - 0.9) | 0.022 | 0.125 | 0.012 |
| *Streptococcus parasanguinis* | 1.2<br>(0.09 - 3.9) | 1.2<br>(0.5 - 1.6) | 0.8<sup>a</sup><br>(0.2 - 2.2) | 0.3<br>(0.09 - 1.4) | 0.6<br>(0.09 - 1.8) | 1.5<sup>b</sup><br>(0.1 - 2.5) | 0.5<br>(0.2 - 1.3) | 0.8<br>(0.09 - 3.6) | 0.5<sup>c</sup><br>(0.1 - 2.3) | 0.780 | 0.746 | 0.018 |
| *Streptococcus pneumoniae* | 13.5<br>(5.5 - 17.3) | 13.1<br>(6.1 - 24.2) | 11.3<br>(6.8 - 20.3) | 12.2<br>(4.8 - 17.4) | 9.6<br>(5.3 - 15.5) | 8.9<br>(5.2 - 14.5) | 10.4<br>(6.8 - 15.5) | 9.5<br>(6.8 - 14.0) | 10.7<br>(6.8 - 21.5) | 0.577 | 0.734 | 0.429 |
| *Streptococcus pyogenes* | 2.9<br>(2.5 - 3.4) | 2.8<br>(2.1 - 3.3) | 2.8<br>(1.4 - 3.1) | 2.8<br>(2.0 - 4.4) | 2.6<br>(2.1 - 3.6) | 2.5<br>(2.1 - 3.6) | 2.4<br>(2.0 - 3.5) | 2.6<br>(1.6 - 3.7) | 2.6<br>(2.3 - 3.4) | 0.948 | 0.363 | 0.727 |
| *Streptococcus salivarius* | 2.9<br>(1.3 - 22.1) | 2.6<br>(1.4 - 11.7) | 2.4<br>(1.0 - 22.8) | 3.1<br>(1.6 - 35.2) | 4.2<br>(2.0 - 34.8) | 12.6<br>(1.3 - 23.5) | 3.8<br>(1.0 - 26.5) | 6.3<br>(1.4 - 12.4) | 10.0<br>(1.7 - 10.6) | 0.570 | 0.758 | 0.713 |
| *Streptococcus sp. HSISS1* | 5.9<br>(1.0 - 12.6) | 7.1<br>(0.1 - 12.7) | 4.3<br>(0.5 - 11.5) | 4.6<br>(0.4 - 12.8) | 4.3<br>(1.3 - 6.5) | 3.8<br>(1.8 - 9.7) | 5.0<br>(2.1 - 10.0) | 4.5<br>(0.3 - 9.4) | 3.5<br>(1.5 - 8.8) | 0.809 | 0.814 | 0.681 |
| *Streptococcus sp.*<br>*LPB0220* | 1.8<br>(0.6 - 10.7) | 3.3<br>(0.9 - 6.8) | 1.3<br>(0.5 - 9.8) | 3.5<br>(0.5 - 13.0) | 5.9<br>(1.4 - 10.8) | 8.0<br>(2.7 - 11.1) | 5.9<br>(0.3 - 12.4) | 0.8<br>(0.1 - 11.3) | 1.6<br>(0.08 - 7.2) | 0.446 | 0.802 | 0.308 |
| *Streptococcus suis* | 3.4<br>(2.0 - 6.3) | 3.5<br>(1.9 - 4.8) | 2.9<br>(1.5 - 4.9) | 3.5<br>(1.6 - 8.4) | 3.6<br>(2.0 - 7.6) | 4.1<br>(1.3 - 6.2) | 2.6<br>(1.4 - 7.0) | 3.5<br>(1.1 - 4.9) | 3.2<br>(1.6 - 4.5) | 0.635 | 0.885 | 0.927 |
| *Streptococcus thermophilus* | 8.9<br>(7.7 - 10.1) | 8.5<br>(7.3 - 10.3) | 8.6<br>(3.9 - 9.5) | 9.6<br>(6.4 - 16.7) | 9.5<br>(6.9 - 16.8) | 9.2<br>(6.4 - 13.5) | 7.4<br>(6.4 - 12.9) | 8.3<br>(4.4 - 10.1) | 8.6<br>(5.6 - 10.2) | 0.401 | 0.329 | 0.243 |
| *Lachnoanaerobaculum umeaense* | 1.4<br>(0.06 - 4.0) | 1.4<br>(0.2 - 3.8) | 0.9<br>(0.1 - 9.0) | 0.1<br>(0.05 - 3.9) | 0.9<br>(0.1 - 1.7) | 0.5<br>(0.03 - 5.4) | 1.1<br>(0.5 - 2.5) | 2.0<br>(0.2 - 5.2) | 1.6<br>(0.2 - 4.6) | 0.458 | 0.620 | 0.868 |
| *Veillonella atypica* | 1.5<br>(0.01 - 10.2) | 1.6<br>(0.02 - 13.7) | 0.3<br>(0.03 - 3.0) | 0.5<br>(0.07 - 3.3) | 0.6<br>(0.6 - 6.2) | 2.5<br>(1.0 - 4.0) | 4.1<br>(0.3 - 10.7) | 1.4<br>(0.1 - 8.4) | 2.8<br>(1.0 - 4.3) | 0.437 | 0.640 | 0.129 |
| *Veillonella nakazawae* | 0.7<br>(0.1 - 1.0) | 0.5<br>(0.1 - 1.7) | 0.3<br>(0.2 - 0.3) | 0.1<br>(0.1 - 0.3) | 0.3<br>(0.2 - 0.5) | 0.3<br>(0.1 - 0.4) | 0.1<br>(0.04 - 0.6) | 0.4<br>(0.4 - 0.4) | 0.05<br>(0.05 - 0.08) | 0.158 | 0.683 | 0.946 |
| *Veillonella parvula* | 6.9<br>(2.8 - 9.6) | 4.8<br>(2.9 - 15.8) | 2.1<sup>a</sup><br>(0.03 - 4.9) | 3.4<br>(1.3 - 3.9) | 5.1<br>(3.1 - 7.1) | 4.7<sup>b</sup><br>(0.5 - 7.9) | 4.6<br>(1.3 - 9.6) | 3.6<br>(0.5 - 8.9) | 2.9<sup>b</sup><br>(1.8 - 5.2) | 0.608 | 0.053 | 0.043 |

Values are presented as median and range. Different letters mean significant differences ( $p < 0.05$ ) and were calculated following a general linear model of covariance followed by the Bonferroni correction for multiple tests.

*Granulicatella elegans* exhibited the greatest variation (p < 0.01). Patients with cancer receiving a standard dose of DMB were found to have a greater presence of that bacteria in their oral microbiome compared with patients receiving a high dose of DMB and placebo. There were substantial changes in two species of *Streptococcus*, *Streptococcus mutans* (p= 0.012) and *Streptococcus parasanguinis* (p= 0.018). For *Streptococcus mutans*, we observed higher abundances in the standard dose DMB group compared to both the high dose of DMB and placebo groups. The levels of *Streptococcus parasanguinis* in the DMB groups exhibited an inverse pattern, with lower abundances in the standard dose DMB group, whereas the abundances in the placebo group remained unchanged. Within the DMB groups, *Veillonella parvula* exhibited opposite variations, with significantly lower abundances in the standard dose DMB group compared with both the high dose DMB and the placebo groups (**Table 2**).

### Microbiome balance

The Rivera-Pinto method [56] was employed to ascertain the microbiome balance at the end of the trial. The analysis revealed that the standard dose DMB group was characterized by lower balance scores for *Streptococcus mutans* and *Bacillus* when compared to *Gemella sp.* zg-570 and *Veillonella* genera (**Figure 1A**). On the contrary, *Gemella sp.* zg-570 and *Veillonella* genera were more associated with the placebo group than with the standard dose DMB group (**Figure 1A**). Concerning the high dose DMB group versus the placebo group, *Gemella sanguinis* and *Streptococcus lutetiensis* were most associated with the placebo group (**Figure 1B**). Thus, at the higher dose of DMB, lower balance scores were associated with lower relative abundances of *Streptococcus intermedius* and *Streptococcus agalactiae* when compared to *Gemella sanguinis* and *Streptococcus lutetiensis* (**Figure 1B**).

**Figure 1.**
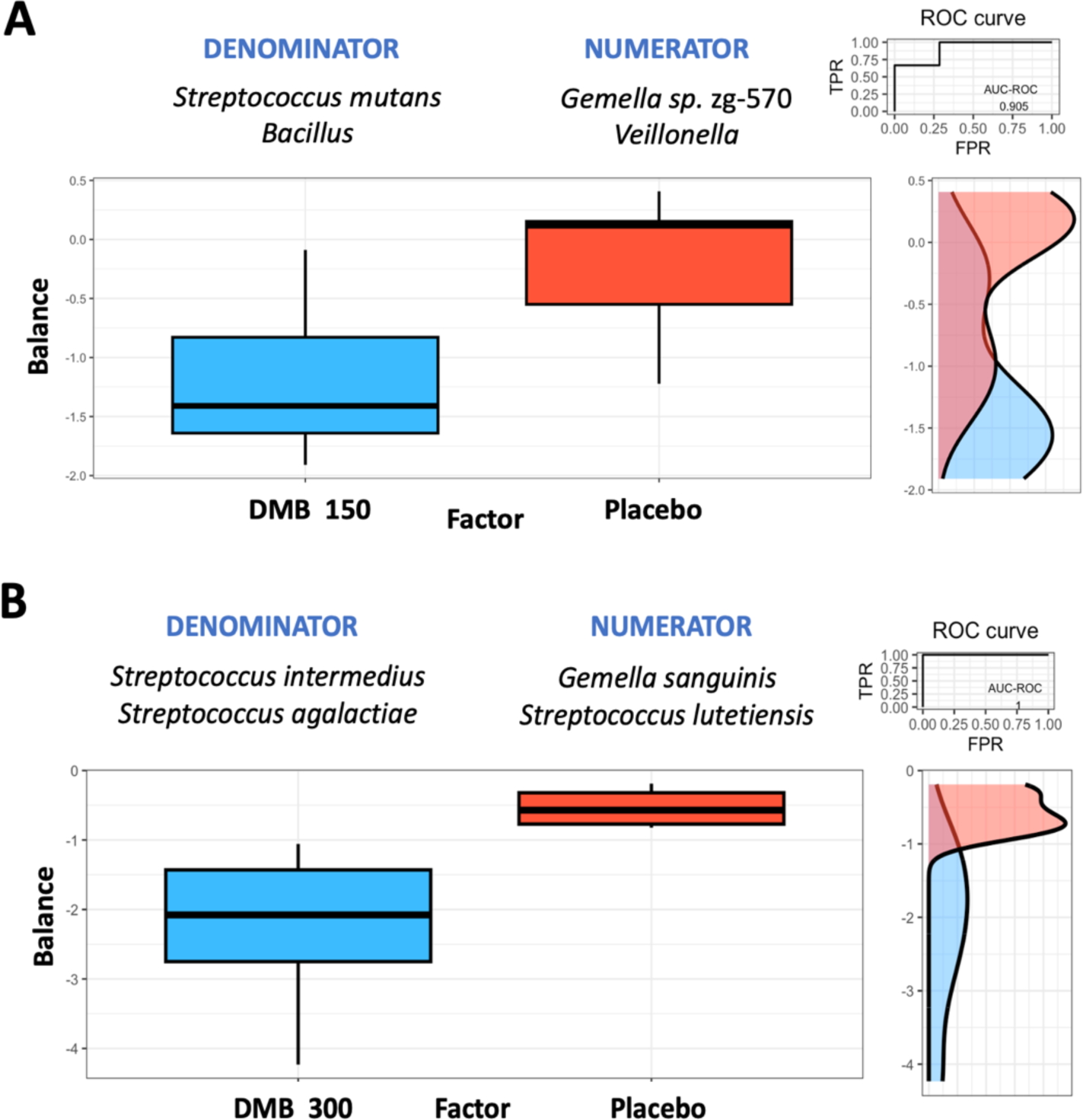
Group microbial balances are presented in an overview. It is indicated at the top of the plot that groups of taxa constitute the global balance. Box plots illustrate the distribution of balance scores for the DMB 150 mg (standard dose) and placebo groups (A) and the DMB 300 mg (high dose) and placebo groups (B). On the right, the ROC curve with its AUC value and the density curve are displayed.

### Analysis of the relationships between oral microbiome, inflammatory cytokines, nutritional status, and electrical taste perception in the three groups

There were significant correlations between the abundance of several species in the standard dose DMB group and a variety of outcomes, as shown in **Figure 2A**. The abundance of *Streptococcus* genus, particularly species of *Streptococcus thermophilus*, *Streptococcus pneumoniae*, *Streptococcus dysgalactiae* and *Streptococcus agalactiae*, correlated negatively with the concentration levels of TNF-α in patients with cancer. Furthermore, energy intake (%) and lipid percentage of energy, as well as monounsaturated fatty acids (MUFAs) and polyunsaturated fatty acids (PUFAs) expressed as percentages of energy were positively correlated with those bacteria. Additionally, MUFAs and PUFAs percentages of energy were positively associated with *Veillonella parvula* levels (**Figure 2A**). The group that received the highest dose of DMB showed several correlations with other outcomes of the study (**Figure 2B**). Indeed, the relative abundance of *Streptococcus pneumoniae*, *Streptococcus dysgalactiae*, and *Streptococcus agalactiae* was positively correlated with energy intake. Saturated fatty acids were positively associated with *Streptococcus pneumoniae* and *Streptococcus dysgalactiae* levels. Lastly, *Granulicatella adiacens* was negatively associated with TNF-α concentration (**Figure 2B**). In the placebo group, the relative abundance of *Granulicatella adiacens* was negatively correlated with the energy intake. The levels of *Streptococcus thermophilus* were negatively correlated with electric taste perception on both the right and left sides. The PUFAs intake was positively associated with *Streptococcus agalactiae* (**Figure 2C**). Neither selected bacteria (*Granulicatella adiacens*, *Streptococcus agalactiae*, *Streptococcus dysgalactiae*, *Streptococcus pneumoniae*, and *Veillonella parvula*) nor electrogustometry, the type of diet, cytokine levels, were associated with mucositis, according to the binary logistic regression. There were only two variables that remained in the equation without reaching statistical significance: TNF-α levels (p=0.109) and saturated fatty acids (p=0.142).

**Figure 2.**
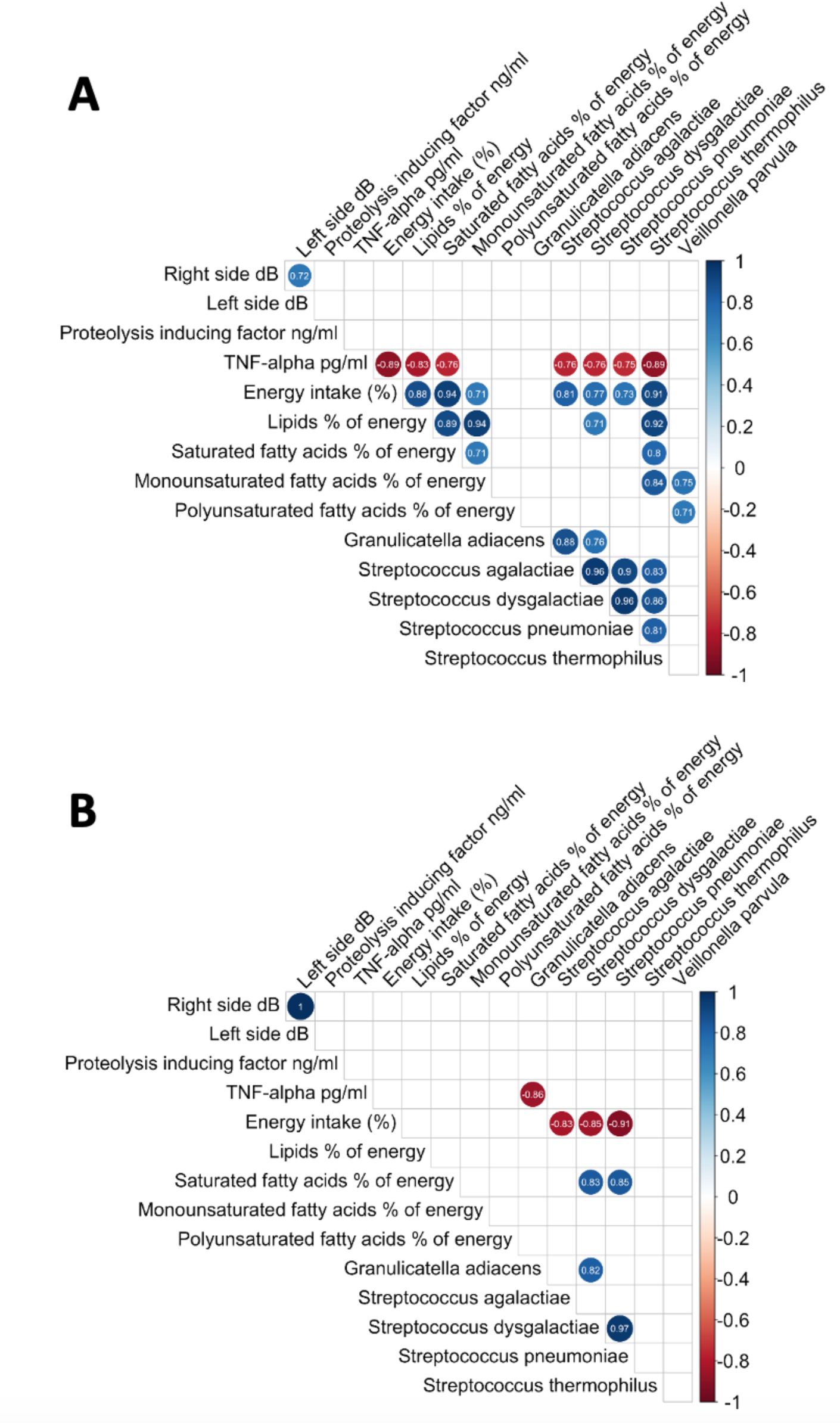
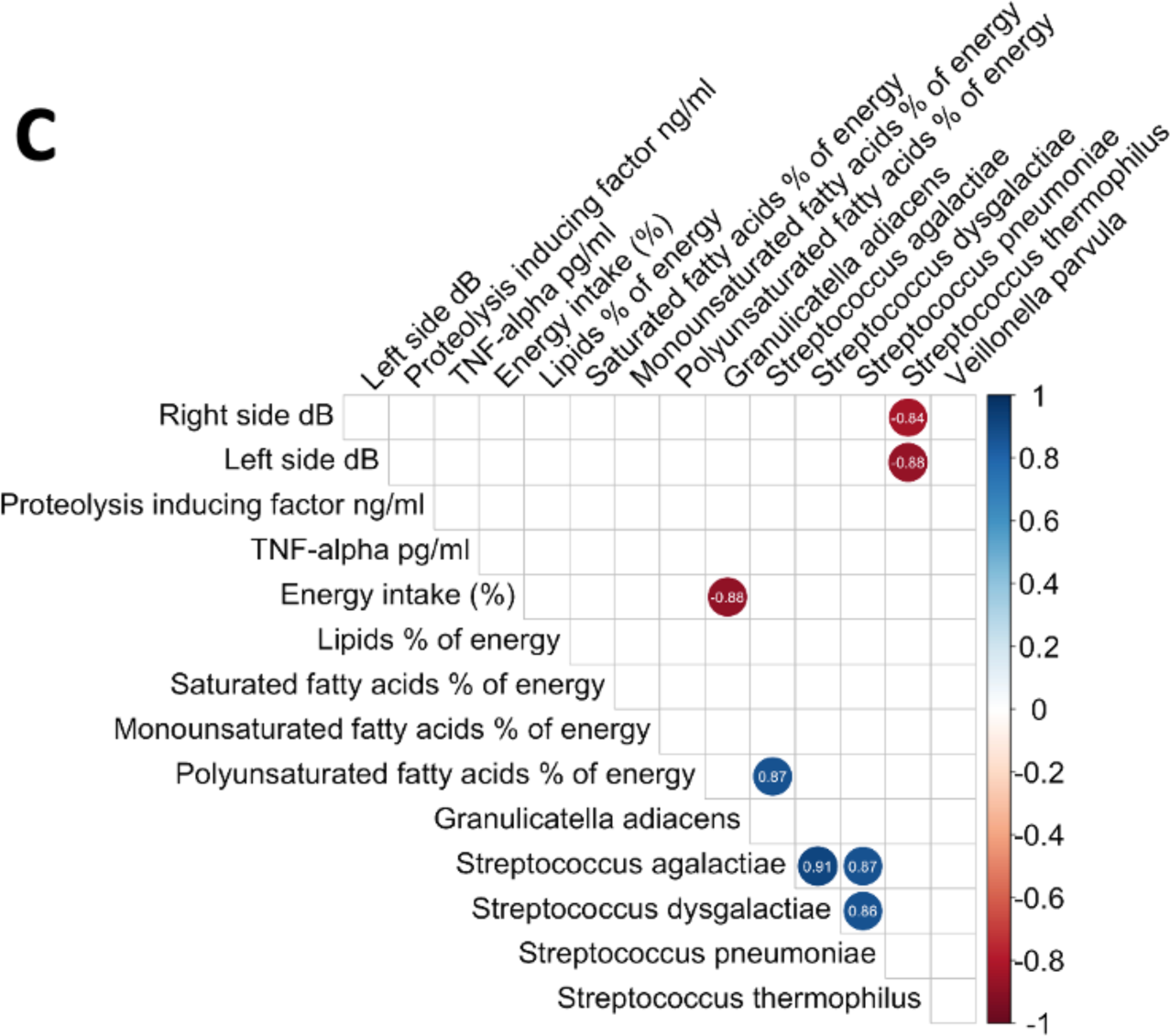
Correlations between the oral microbiome, inflammatory cytokines, nutritional status, and electrical taste perception. **A.** DMB 150 mg (standard dose), **B.** DMB 300 mg (high dose), and **C.** placebo.

Moreover, no independent correlation was found between the presence of selected species and clinical outcomes evaluated as immunotherapeutic agents used for the cancer treatment, general toxicity, digestive toxicity, or consumption of tobacco and alcohol.

## Discussion

In the present study, patients with cancer, malnourished and with dysgeusia presented dysbiosis. We have shown that regular consumption of 150 mg of DMB (standard dose), as an adjuvant to medical-nutritional treatment, changed the oral microbiome composition in these patients receiving antineoplastic treatment. In particular, the consumption of the standard dose of DMB resulted in a greater relative abundance of *Enterococcus*, and a lower abundance of the *Veillonella* genus that did the consumption of a high dose of DMB or placebo. Additionally, some species such as *Granulicatella elegans*, *Granulicatella adiacens Streptococcus mutans*, and *Gemella morbillorum* exhibited greater relative abundances in patients receiving the standard dose of DMB. However, lower abundances of *Streptococcus parasanguinis*, *Veillonella parvula*, *Streptococcus australis* and *Streptococcus cristatus* were detected. The consumption of the standard dose of DMB revealed a link between several *Streptococcus* species and lower TNF-α plasma levels, as well as higher energy and plasma MUFAs and PUFAs dietary intake.

The oral microbiota is composed of a multitude of bacterial species, including *Actinomycetota* (formerly *Actinobacteria*), *Bacteroidota* (*Bacteroidetes*), *Bacillota* (*Firmicutes*), *Fusobacteriota* (*Fusobacteria*), *Pseudomonadota* (*Proteobacteria*), *Saccharibacteria* and *Spirochaetota* (*Spirochaetes*) [24]. These bacterial species are generally conserved across individuals, making up the bulk of the oral microbiota [58], which plays a relevant role in oral health [59]. The mechanism by which commensals promote healthy oral microbiota involves outcompeting pathogens for colonization [60]. Microbes temporarily overpower the immune system in the event of reduced diversity and richness, described as dysbiosis [61, 62]. Nevertheless, the oral microbiome serves more than just a local function, as microbes are capable of communicating with the entire body [63]. The presence of oral dysbiosis can contribute to the maintenance of chronic low-grade systemic inflammation by causing a localized inflammatory state within the oral cavity [64].

Cancer can be affected by microbes in a variety of ways, including contact-dependent effects occurring locally at the mucosal surface or within the tumor microenvironment. The second type of effect is contact-independent effects, which are caused by the metabolites produced by microbes and the vesicles of their outer membranes that circulate in the blood [65, 66]. Thus, several types of cancer may be associated with specific patterns of salivary and fecal microbiomes, as well as circulating microbial DNA in blood plasma [67].

Alterations in the oral microbiome that are associated with cancer treatments cause dysbiosis, which is a condition characterized by an imbalanced status of the oral microbial community [68]. Thus, the dysbiosis of the oral microbiome is characterized by an increase in the prevalence of pathogenic microbial species at the expense of commensal microorganisms and a markedly reduced richness and diversity of species [69, 70]. Taste disorders are frequently reported by oncologic patients undergoing antineoplastic treatments [3, 4]. Consequently, taste disorders have a significant impact on eating behaviors and, as a result, have a major influence on overall health [59]. In addition, this alteration might cause oral microbiome dysbiosis during cancer therapies, characterized by a markedly reduced richness and diversity of species [70]. Here, we observed that the richness and diversity, measured by the Shannon and Simpson indices, along with the Chao1 index, were reduced in our patients with cancer compared to the levels reported in healthy individuals [57], indicating basal dysbiosis in all groups.

*Gemella* was the fourth most prevalent genera in our study, followed by *Streptococcus*, *Veillonella*, and *Granulicatella*. *Gemella sanguinis* was the most prevalent *Gemella* species in our study. A significantly greater enrichment of the *Gemella* genus in oral squamous cell carcinoma has been reported. In oral leukoplakia and oral squamous cell carcinoma, *Streptococcus* sp. NPS 308, *Streptococcus agalactiae*, *Gemella hemolysans*, and *Gemella morbillorum* were slightly increased [71]. A high relative abundance of *Gemella morbillorum* is present in oral cavity squamous cell carcinoma tumor tissues compared with the paired adjacent normal tissues [72]. Here, we observed a tendency for *Gemella morbillorum* to be more abundant in the standard dose of DMB group; however, lower levels were observed in the high dose of DMB group, indicating a different profile dose-dependent for this bacterium. *In vitro* studies with three oral squamous cell carcinoma cell lines (CAL27, SCC4, and SCC25), have demonstrated the positive effects of oral commensals belonging to the *Streptococcus* genus [73]. In our patients with cancer, these species make up more than 63% of the total oral microbiome. According to the above data, these bacteria are capable of acting as anticancer agents.

*Veillonella parvula* is elevated in oral cancer [74], especially in oral squamous cell carcinoma, by promoting the expression of inflammatory cytokines, including interleukins (IL-6, IL-8) and TNF-α [75]. Here, we observed that regular consumption of a standard dose of DMB decreases the abundance of *Veillonella parvula*, an important oncologic-related bacteria [75]. Hence, the lower relative abundance of this bacteria in the standard dose DMB group might be linked to the improvement in systemic inflammation and tumor markers observed in malnourished patients with cancer.

At the species level, we also found that the relative abundance of *Granulicatella elegan*s, *Granulicatella adiacens*, and *Streptococcus mutans* were greater in patients receiving the standard dose of DMB. However, lower abundances of *Streptococcus parasanguinis*, *Streptococcus australis* and *Streptococcus cristatus* were detected.

Regarding the genus *Granulicatella*, we observed a significant variation in the abundance of *Granulicatella elegans*, with an increase in the standard dose DMB group, and a decrease in the high dose DMB group. This is of interest since a positive association has been found between the abundance of *Granulicatella elegans* and inflammation [76]. In the case of *Granulicatella adiacens*, higher levels at both standard and high doses of DMB were observed. Microbiome-associated pathology can arise from changes in general bacterial composition, such as those found in periodontitis, and from colonization and overgrowth of keystone species. The infiltration of immunosuppressive cells and the interference of immune killer cells by commonly occurring oral microorganisms, such as *Fusobacterium nucleatum* and *Porphyromonas gingivalis*, can prevent tumor cells from being observed and cleared by the immune system [77–79]. The absence of periodontitis and gingivitis in these patients with cancer may explain our failure to detect any of these bacteria in our study. There is evidence that a high abundance of *Fusobacterium nucleatum* in the oral microbiome of patients with colorectal cancer is associated with tumor metastasis, recurrence, chemo-resistant cancer, and reduced radiotherapy efficacy [80–83].

Further, in our study, neither of the highly represented bacteria was associated with the presence or absence of mucositis at the end of the study based on binary logistic regression. It is also important to note that no independent correlation was found between the presence of selected species and clinical outcomes evaluated as immunotherapeutic agents used for the cancer treatment, general toxicity, digestive toxicity, or consumption of tobacco and alcohol.

The oral *Streptococcus* genus plays a key role in oral dysbiosis and a wide range of clinical conditions, including dental caries, gingivitis, periodontal disease, and oral cancer [84]. We found that *Streptococcus thermophilus*, *Streptococcus pneumoniae*, *Streptococcus dysgalactiae*, and *Streptococcus agalactiae* were negatively associated with the plasma levels of TNF-α in malnourished patients with cancer receiving a standard dose of DMB, which might be associated with the overall improvement of systemic inflammation after DMB supplementation. There are some strains of *Streptococcus* that have inherent antitumor activity or that can activate the immune system of the host to fight tumors [85]. In addition, those bacterial species were positively associated with energy intake, lipid percentage of energy, plasma MUFAs and PUFAs, expressed as percentages of energy. Moreover, the percentages of energy-related MUFAs and PUFAs were positively associated with *Veillonella parvula* levels. This finding is of interest since we observed an improvement in quality of life in malnourished patients with cancer who received a standard dose of DMB [41]. Therefore, oral intake of DMB might improve the pattern of oral bacteria and in turn, reduce inflammation. In the case of the group that received high doses of DMB, we observed that the relative abundances of *Streptococcus pneumoniae*, *Streptococcus dysgalactiae*, and *Streptococcus agalactiae* were positively correlated with energy intake. Saturated fatty acids were positively associated with *Streptococcus pneumoniae* and *Streptococcus dysgalactiae* levels. The abundance of *Granulicatella adiacens* was negatively associated with the plasma TNF-α concentration, indicating an improvement in the systemic inflammation associated with oral dysbiosis. Finally, there was no correlation between changes in the oral microbiome of the DMB groups and electric taste perception. It was only in the placebo group that *Streptococcus thermophilus* abundance was negatively correlated with electric taste perception.

Besides, *Streptococcus mutans* is involved in the etiology of several oral diseases [86–88]. In particular, *Streptococcus mutans* is implicated as the main etiologic agent of caries [89–91]. Dysbiosis of the dental plaque microbiome is associated with an abundance of biofilm-forming, acid-producing, and acid-tolerant species. Consumption of the standard dose of DMB was associated with a greater relative abundance of *Streptococcus mutans*. This might be linked to higher energy consumption by those patients. However, a lower relative abundance of *Streptococcus mutans* was observed in patients receiving high doses of DMB. Nevertheless, compared with patients in the standard dose group these patients did not show an improvement in quality of life [41].

While *Streptococcus mutans* is associated with caries, *Streptococcus cristatus* plays a relevant role in the development of periodontitis, a common oral disease and it may contribute to the pathogenicity of the oral microbiome [92, 93]. Here, we observed a tendency to decrease the relative abundance of *Streptococcus cristatus* in the standard dose DMB group compared to both the high dose DMB group and the placebo group.

Overall, the changes of the oral microbiota observed in patients having the standard dose of DMB differed compared with those having the higher dose. This could be due to the fact that the latter group manifest to have a sweet taste for much time after the consumption of the DMB tablet, compared to the former, which in turn results into a lower dietary intake [41].

### Strengths and limitations

To the best of our knowledge, this is the first attempt to evaluate the potential effects of DMB on patients with cancer and dysgeusia. The patients in the pilot study followed their usual nutritional and medical recommendations. Additional nutritional and medical monitoring were provided as part of their participation in the CLINMIR study. The DMB was added as an adjuvant to medical-nutritional treatment. The major strength of the present study is that we used a randomized triple-blind placebo-controlled study for the evaluation of the effects of DMB on the oral microbiome. Additionally, the use of the Nanopore technology, which allows the whole sequence of the 16S RNA gene and the identification of microbiome at the species level compared with other techniques that only allow the identification at the genus level, is also one major strength of this study. However, this study has certain limitations; first, the number of participants was insufficient to provide a robust basis for the conclusions drawn from the data. Moreover, we performed our analysis considering missing patient data. Consequently, the outcomes may be limited, and future investigations utilizing a larger cohort would be invaluable in elucidating the potential benefits of DMB consumption for patients with cancer.

### Conclusions

To identify innovative therapies for the treatment of taste disorders in patients with cancer, we conducted this pilot randomized, parallel, triple-blind, and placebo-controlled clinical trial aimed at providing a novel strategy for reducing the side effects of chemotherapy, radiotherapy, and immunotherapy, including alterations in taste and changes in body composition, nutritional status, and quality of life [41]. All patients presented dysbiosis in terms of bacterial diversity and richness, compared with healthy individuals. Here, we showed that regular consumption of a standard dose of DMB, as an adjuvant to medical-nutritional treatment, seems to improve the oral microbiome composition in malnourished patients with cancer receiving antineoplastic treatment. Thus, we reported that the relative abundances of *Granulicatella elegans*, *Granulicatella adiacens, Streptococcus mutans*, and *Gemella morbillorum* were greater in patients with cancer receiving a standard dose of DMB. However, lower abundances of *Streptococcus parasanguinis*, *Veillonella parvula*, *Streptococcus australis* and *Streptococcus cristatus* were detected. Furthermore, DMB consumption revealed a negative association between *Streptococcus thermophilus*, *Streptococcus pneumoniae*, *Streptococcus dysgalactiae*, and *Streptococcus agalactiae* and TNF-α concentrations in patients with cancer. The presence of *Streptococcus thermophilus* and *Veillonella parvula* was positively associated with plasma MUFAs, with only *Veillonella parvula* being associated with plasma PUFAs. Overall, DMB intake improves the oral microbiome in patients with cancer and dysgeusia, which may contribute to a better immune response.

## Data Availability Statement

A reasonable request should be made to the corresponding author for access to the datasets used and/or analyzed in the current study.

## Funding statement

This study is funded by Medicinal Gardens S.L. through the Center for Industrial Technological Development (CDTI), “Cervera” Transfer R&D Projects. Ref. IDI-20210622. (Science and Education Ministry, Spain).

## Conflicts of Interest

The authors declare that they have no commercial or financial relationships that could be construed as potential conflicts of interest.

## Ethics approval statement

The study was conducted under the Declaration of Helsinki, and approved by the Ethics Committee of La Paz University Hospital (protocol code 6164, 23 June 2022).

## Patient consent statement

Patient informed consent was obtained from all subjects involved in the study.

## Clinical trial registration

The CLINMIR study is a pilot randomized, parallel, triple-blind, and placebo-controlled clinical trial. Using the number NCT05486260, the present protocol was registered at http://clinicaltrials.gov, accessed on 25 June 2024.

## Author Contributions

Conceptualization, B.L.-P., A.G. and S.P.-M.; methodology, B.L.-P. and J.F.-B.; software, J.D.-P.; validation, F.J.R.-O, A.I.A.-M. and M.B.-H.; formal data analysis, J.D.-P., F.J.R.-O, and M.B.-H.; investigation, B.L.-P and L.A.-C.; resources, S.P.-M.; data curation, L.A.-C; writing-original draft preparation, J.D.-P., F.J.R.-O, M.B.-H. and A.G.; writing-review and editing, J.D.-P., F.J.R.-O, M.B.-H. and A.G.; supervision, S.P.-M and A.G.; project administration, B.L.-P; funding acquisition, S.P.-M. All authors have read and agreed to the published version of the manuscript.

## Acknowledgments

J.P.-D. is part of the “UGR Plan Propio de Investigación 2016” and the “Excellence Actions: Unit of Excellence in Exercise and Health (UCEES), University of Granada”. F.J.R.-O. is supported by a grant from the Spanish Government’s “Agencia Estatal de Investigación-Juan de la Cierva-Incorporación” program (IJC2020-042739-I). We thank (Lucía Tadeo, Helena Torrell, Adría Cereto and Núria Canela) from the Genomics facility of the Centre for Omic Sciences (COS) Joint Unit of the Universitat Rovira i Virgili-Eurecat, for their contribution to sequencing analysis.

## Notes

### Competing Interest Statement

The authors have declared no competing interest.

### Clinical Trial

NCT05486260

### Clinical Protocols

https://clinicaltrials.gov/study/NCT05486260?tab=results

