## Supplementary material for "Effect of a novel food rich in miraculin on the oral microbiome of malnourished oncologic patients with dysgeusia": Tables S1-S3

**Table S1**. Nutritional composition of the food supplement enriched in miraculin (DMB) and placebo

| | | Standard dose of DMB (150 mg DMB + 150 mg strawberry freeze-dried) | High dose of DMB (300 mg DMB) | Placebo (300 mg strawberry freeze-dried) |
|---|---|---|---|---|
| Energy | kcal | 0.99 | 1 | 0.97 |
| Carbohydrates | mg | 194 | 234 | 154 |
| Sugars | mg | 156 | 162 | 150 |
| Fiber | mg | 26 | 6 | 46 |
| Proteins | mg | 20 | 15 | 24 |
| Lipids | mg | 9 | 5 | 12 |
| Saturated fatty acids | mg | 2 | 2 | 1 |
| Sodium chloride | mg | 0.1 | 0.1 | 0.03 |
| Humidity | mg | 4 | 4 | 5 |
| Ash | mg | 12 | 14 | 15 |
| Miraculin | mg | 2,8 | 5,6 | 0 |

Nutritional composition provided by Medicinal Gardens, S.L.

**Table S2.** Distribution of phyla and alpha diversity indices for microbiota of saliva from cancer patients of the CLINMIR study.

| Phylum | Standard Dose DMB (150 mg) | | | High Dose DMB (300 mg) | | | Placebo | | | p-value | | |
|---|---|---|---|---|---|---|---|---|---|---|---|---|
| | Baseline | 1 month | 3 months | Baseline | 1 month | 3 months | Baseline | 1 month | 3 months | Treatment (T) | Time (t) | T x t |
| *Actinobacteriota* | 0.6 (0.2 - 2.4) | 0.5 (0.09 - 2.0) | 0.5 (0.2 - 2.6) | 0.7 (0.2 - 0.9) | 0.5 (0.4 0.8) | 0.4 (0.2 - 0.5) | 0.6 (0.2 - 1.4) | 0.7 (0.2 - 2.3) | 0.6 (0.2 - 1.6) | 0.469 | 0.556 | 0.926 |
| *Bacteroidota* | 0.01 (0.005 - 0.02) | 0.01 (0.007 - 0.02) | 0.03 (0.01 - 0.05) | 0.01 (0.005 - 0.03) | 0.01 (0.009 - 0.06) | 0.01 (0.01 - 0.01) | 0.01 (0.004 - 0.02) | 0.01 (0.007 - 0.06) | 0.01 (0.005 - 0.01) | 0.815 | 0.119 | 0.180 |
| *Bacillota* | 99.2 (97.0 - 99.6) | 99.1 (97.6 - 99.7) | 99.2 (96.6 - 99.7) | 99 (98.2 - 99.8) | 99.3 (98.7 - 100) | 99.5 (99 - 99.6) | 98.9 (98.1 - 99.7) | 98.9 (95.1 - 99.3) | 98.9 (98.3 - 99.4) | 0.254 | 0.630 | 0.620 |
| *Fusobacteria* | 0.02 (0.004 - 0.03) | 0.01 (0.004 - 0.05) | 0.03 (0.004 - 0.06) | 0.01 (0.01 - 0.01) | 0.01 (0.01 - 0.01) | 0.01 (0.006 - 0.05) | 0.006 (0.004 - 0.04) | 0.01 (0.003 - 0.04) | 0.007 (0.007 - 0.007) | 0.521 | 0.619 | 0.905 |
| *Pseudomonadota* | 0.1 (0.05 - 0.3) | 0.1 (0.06 - 0.4) | 0.12 (0.05 - 0.5) | 0.06 (0.03 - 0.3) | 0.07 (0.03 - 0.2) | 0.07 (0.04 - 0.4) | 0.1 (0.06 - 0.2) | 0.2 (0.08 - 0.4) | 0.2 (0.08 - 0.3) | 0.534 | 0.343 | 0.187 |
| *Saccharibacteria* | 0.04 (0.01 - 0.4) | 0.08 (0.01 - 0.3) | 0.05 (0.008 - 0.4) | 0.05 (0.02 - 0.3) | 0.07 (0.05 - 0.3) | 0.06 (0.01 - 0.1) | 0.2 (0.004 - 0.4) | 0.1 (0.03 - 2) | 0.1 (0.05 - 0.3) | 0.252 | 0.310 | 0.391 |
| *Shannon index* | 1.4 (1.2 - 1.5) | 1.5 (1.1 - 2.0) | 1.4 (1.1 - 2.6) | 1.2 (0.4 - 2.1) | 0.8 (0.6 - 1.7) | 1.3 (0.8 - 1.7) | 1.5 (0.8 - 1.9) | 1.6 (1.1 - 2.6) | 1.4 (1.1 - 2.2) | 0.516 | 0.373 | 0.591 |
| *Simpson's index* | 0.5 (0.5 - 0.6) | 0.6 (0.4 - 0.7) | 0.5 (0.4 - 0.8) | 0.5 (0.1 - 0.7) | 0.4 (0.3 - 0.6) | 0.5 (0.3 - 0.7) | 0.6 (0.3 - 0.7) | 0.6 (0.4 - 0.9) | 0.5 (0.5 - 0.8) | 0.673 | 0.428 | 0.714 |
| *Chao1 index* | 41.0 (22.8 - 60.0) | 39.9 (32.0 - 57.2) | 38.0 (24.3 - 64.0) | 37.5 (21.0 - 47.0) | 32.4 (4.0 - 79.0) | 38.6 (30.2 - 52.0) | 36.2 (22.0 - 49.0) | 39.6 (24.8 - 66.0) | 41.5 (23.8 - 54.7) | 0.575 | 0.411 | 0.405 |

Values are presented as median and range. General linear mixed models (GLM) of variance (ANOVA) were used to evaluate differences between means for treatment, time, and treatment x time.

**Table S3.** Distribution of selected families for microbiota of saliva from cancer patients of the CLINMIR study.

| | Standard Dose DMB (150 mg) | | | High Dose DMB (300 mg) | | | Placebo | | | p-value | | |
|---|---|---|---|---|---|---|---|---|---|---|---|---|
| Selected families | Baseline | 1 month | 3 months | Baseline | 1 month | 3 months | Baseline | 1 month | 3 months | Treatment (T) | Time (t) | T x t |
| *Streptococcaceae* | 69.4 (61.3 - 71.2) | 64.3 (56.5 - 75.2) | 68.7 (37.1 - 77.2) | 72.0 (53.0 - 94.0) | 74.4 (60.0 - 84.5) | 69.0 (53.0 - 84.9) | 64.9 (49.6 - 85.2) | 63.0 (33.6 - 76.8) | 68.0 (41.2 - 72.5) | 0.333 | 0.545 | 0.586 |
| *Veillonellaceae* | 10.5 (3.6 - 20.9) | 6.9 (3.5 - 27.8) | 3.9 (0.07 - 6.0) | 5.3 (1.7 - 8.1) | 6.8 (4.6 - 15.8) | 7.4 (3.5 - 15.5) | 11.0 (3.1 - 24.4) | 6.5 (2.9 - 14.0) | 5.9 (4.6 - 8.4) | 0.436 | 0.856 | 0.906 |
| *Carnobacteriaceae* | 4.9 (1.7 - 14.7) | 3.7 (2.2 - 8.0) | 4.9 (1.4 - 9.2) | 1.6 (0.09 - 7.4) | 1.6 (0.3 - 4.8) | 4.0 (0.8 - 23.3) | 3.0 (1.4 - 8.8) | 4.8 (1.3 - 7.4) | 2.7 (1.3 - 3.2) | 0.733 | 0.463 | 0.936 |
| *Lachnospiraceae* | 3.8 (1.5 - 5.2) | 5.4 (1.2 - 10.2) | 4.2 (1.4 - 27.0) | 1.5 (0.08 - 17.9) | 4.8 (0.5 - 6.9) | 2.5 (0.2 - 18.9) | 3.6 (2.4 - 8.4) | 8.2 (1.7 - 15.9) | 4.9 (0.9 - 15.4) | 0.128 | 0.312 | 0.345 |
| *Aerococcaceae* | 3.5 (0.2 - 7.9) | 3.1 (0.4 - 14.3) | 0.8 (0.03 - 15.0) | 1.4 (0.3 - 8.5) | 2.0 (1.3 - 6.8) | 1.3 (0.2 - 4.8) | 2.0 (0.1 - 8.4) | 2.4 (0.2 - 11.9) | 0.6 (0.2 - 14.3) | 0.793 | 0.734 | 0.870 |
| *Lactobacillaceae* | 0.9 (0.1 - 13.5) | 1.3 (0.1 - 17.7) | 3.7 (0.2 - 12.9) | 2.3 (0.7 - 7.0) | 0.5 (0.4 - 8.7) | 2.5 (0.3 - 11.9) | 2.4 (0.08 - 12.9) | 2.3 (0.1 - 13.3) | 0.4 (0.1 - 9.0) | 0.124 | 0.312 | 0.162 |
| *Bacillaceae* | 0.9 (0.4 - 2.1) | 0.7 (0.5 - 1.3) | 0.9 (0.6 - 1.6) | 0.3 (0.1- 1.2) | 0.4 (0.2 - 0.6) | 0.7 (0.3 - 3.2) | 0.6 (0.3 - 1.4) | 0.8 (0.4 - 0.9) | 0.5 (0.3 - 0.6) | 0.478 | 0.395 | 0.764 |
| *Eubacteriales Family XIII. Incertae Sedis* | 0.6 (0.07 - 1.5) | 0.5 (0.08 - 2.5) | 0.7 (0.2 - 6.8) | 0.5 (0.05 - 1.1) | 0.4 (0.2 - 0.9) | 0.4 (0.1 - 0.7) | 1.0 (0.2 - 3.6) | 0.7 (0.06 - 5.9) | 0.8 (0.03 - 8.7) | 0.405 | 0.373 | 0.682 |
| *Enterococcaceae* | 0.5 (0.2 - 1.4) | 0.4 (0.3 - 0.8) | 0.5 (0.4 - 0.9) | 2.6 (0.03 - 12.1) | 0.1 (0.04 - 5.8) | 0.4 (0.08 - 2.4) | 0.3 (0.1 - 0.8) | 0.5 (0.2 - 0.6) | 0.3 (0.2 - 11.4) | 0.187 | 0.489 | 0.174 |
| *Listeriaceae* | 0.5 (0.1 - 1.1) | 0.3 (0.2 - 0.6) | 0.6 (0.3 - 0.8) | 0.09 (0.01 - 0.7) | 0.2 (0.06 - 0.3) | 0.3 (0.08 - 1.8) | 0.2 (0.08 - 0.7) | 0.3 (0.1 - 0.4) | 0.2 (0.07 - 0.2) | 0.445 | 0.373 | 0.623 |
| *Actinomycetaceae* | 0.3 (0.1 - 1.2) | 0.4 (0.05 - 1.1) | 0.2 (0.05 - 0.6) | 0.2 (0.08 - 0.5) | 0.3 (0.07 - 0.4) | 0.2 (0.1 - 0.3) | 0.4 (0.2 - 1.3) | 0.4 (0.1 - 1.3) | 0.3 (0.1 - 0.7) | 0.961 | 0.413 | 0.481 |
| *Micrococcaceae* | 0.2 (0.01 - 0.3) | 0.06 (0.01 - 0.3) | 0.09 (0.04 - 0.3) | 0.07 (0.02 - 0.5) | 0.05 (0.02 - 0.1) | 0.03 (0.009 - 0.1) | 0.09 (0.02 - 0.3) | 0.2 (0.02 - 0.3) | 0.07 (0.01 - 0.7) | 0.110 | 0.286 | 0.161 |
| *Staphylococcaceae* | 0.2 | 0.2 | 0.3 | 0.1 | 0.1 | 0.3 | 0.2 | 0.3 | 0.1 | 0.629 | 0.734 | 0.821 |

| | | | | | | | | | | | | |
|---|---|---|---|---|---|---|---|---|---|---|---|---|
| | (0.1 - 0.5) | (0.1 - 0.4) | (0.2 - 0.4) | (0.03 - 0.5) | (0.07 - 0.2) | (0.06 - 0.9) | (0.07 - 0.4) | (0.2 - 1.6) | (0.1 - 0.3) | | | |
| *Clostridiaceae* | 0.1 | 0.1 | 0.2 | 0.08 | 0.1 | 0.08 | 0.2 | 0.2 | 0.2 | 0.508 | 0.338 | 0.847 |
| | (0.04 - 0.2) | (0.03 - 0.4) | (0.03 - 0.5) | (0.02 - 0.4) | (0.04 - 0.3) | (0.02 - 0.5) | (0.04 - 0.7) | (0.03 - 1.2) | (0.02 - 1.4) | | | |

Values are presented as median and range. General linear mixed models (GLM) of variance (ANOVA) were used to evaluate differences between means for treatment, time, and treatment x time.
